# Geographic spillover of antimicrobial resistance from mass distribution of azithromycin

**DOI:** 10.1101/2025.07.22.25331994

**Authors:** Ariktha Srivathsan, Ahmed M. Arzika, Ramatou Maliki, Amza Abdou, Marc Lipsitch, Seth Blumberg, Kieran S. O’Brien, Travis C. Porco, Armin Hinterwirth, Thuy Doan, Jeremy D. Keenan, Thomas M. Lietman, Benjamin F. Arnold

**Affiliations:** Francis I Proctor Foundation, University of California San Francisco, USA; Department of Epidemiology & Biostatistics, University of California San Francisco, USA; Centre de Recherche et Interventions en Santé Publique, Birni N’Gaoure, Niger; Programme National de Santé Oculaire, Niamey, Niger; Center for Communicable Disease Dynamics, Department of Epidemiology, Harvard T.H. Chan School of Public Health, USA; Department of Immunology and Infectious Diseases, Harvard T.H. Chan School of Public Health, USA; Department of Medicine, University of California San Francisco, USA; Department of Ophthalmology, University of California San Francisco, USA; Institute for Global Health Sciences, University of California San Francisco, USA

## Abstract

Large-scale, placebo-controlled, cluster-randomized trials in high-mortality settings in several African countries demonstrated a 14-18% reduction in childhood mortality following twice-annual mass drug administration (MDA) of azithromycin among children aged 1–59 months [1–3]. Azithromycin MDA also selects for antimicrobial resistance (AMR), particularly macrolide resistance in treated populations [4–6]. It is unknown whether the genetic selection of AMR from azithromycin MDA could spill over to neighboring untreated populations. If present, such geographic spillover effects could lead the trials to underestimate the risks of AMR selection from azithromycin MDA. Here, we assessed between-village geographic spillover effects of genotypic resistance to macrolides and other antibiotic classes in rectal swabs collected from 1200 children in 30 monitoring villages in Niger after two years of MDA in 594 surrounding villages. We found no evidence of geographic spillover of macrolide resistance in untreated villages, as the genetic load of AMR remained at baseline levels in placebo-treated villages regardless of surrounding azithromycin treatment intensity. Sensitivity analyses confirmed the robustness of findings to the metric used to quantify the effect of proximal azithromycin MDAs on macrolide AMR, and no geographic spillover effects were detected for AMR to other antibiotic classes. Our results suggest that azithromycin MDA-induced selection of macrolide AMR is localized to treated villages without extending to children in neighboring, untreated villages, mitigating some concerns about geographic spillover of AMR to untreated populations. This analysis illustrates the value of randomized trial designs in assessing indirect effects of large-scale public health interventions.

## Introduction

In 2018 the MORDOR (Macrolides Oraux pour Réduire les Décès avec un Oeil sur la Résistance) trial demonstrated a 14% reduction in all-cause mortality among children 1-59 months following twice-annual mass drug administration (MDA) with azithromycin [1]. This evidence resulted in the World Health Organization (WHO) issuing conditional recommendations that azithromycin MDA be considered for children in high-mortality settings, but the WHO limited the recommendation to children aged 1 to 11 months to help mitigate the potential for selection of antimicrobial resistance (AMR) from MDA [7]. Subsequent trials in Burkina Faso [2] and Niger [3] demonstrated mortality reduction when twice-annual MDA is delivered to all children 1-59 months, but not if delivery was limited to children 1-11 months [3] or if delivered to individual children postnatally [8] or at well-child visits [9]. Based on this evidence, the government of Niger is now considering national-scale, biannual azithromycin MDA to reduce child mortality [10].

Repeated azithromycin MDA to children 1-59 months selects for AMR, particularly to macrolides. Despite the evidence of increased macrolide resistance within treated populations [4–6], it remains unclear whether selection of macrolide resistance could extend to larger spatial scales, potentially transmitting AMR genes to neighboring, untreated populations through person-to-person transmission of resistant bacteria or, potentially, through environmentally mediated transmission. Such “spillover” or indirect effects [11] of AMR could potentially magnify the risks beyond what is currently known [12].

Standard analyses of randomized controlled trials assume independence between randomized units (participants or clusters) [11]. Independence implies that no interference occurs between treated and untreated groups such that the outcome of a randomized unit depends solely on its own treatment, unaffected by the treatment administered to other units [13]. In azithromycin MDA trials, if between-cluster spillover effects are present, the independence assumption does not hold, and the trials could underestimate potential harms: AMR selected in treated villages could spread to untreated villages, making the two groups look more similar than they would in the absence of spillover. Infectious disease intervention trials have quantified such spillover effects across various contexts [11,14–16]. Evidence from HIV prevention [17] and antibiotic resistance to malaria control [18] suggests that even limited interaction between populations can substantially impact intervention outcomes and resistance patterns, with effects observed up to several kilometers away from intervention areas. Notably, phylogenetic analyses of HIV transmission patterns demonstrate that population mobility and geographic proximity are fundamental drivers of spread between neighboring communities [19]. Understanding whether azithromycin MDA can lead to geographic spillover of AMR is crucial for assessing the broader public health implications and potential risks as countries consider scaling up azithromycin MDA to millions of children.

We conducted a pre-specified secondary analysis of the MORDOR trial in Niger, focusing on the potential between-village spillover effects of azithromycin MDA on AMR to macrolides and other antibiotic classes. We hypothesized that if geographic spillover effects were present, placebo-treated communities surrounded by high levels of azithromycin MDA would have a higher genetic load of AMR compared with placebo-treated communities without nearby azithromycin MDA. To test this hypothesis, we leveraged the randomized geographic variation in the distribution of azithromycin doses across 594 villages included in the trial.

## Results

### MORDOR trial: study design and study population

MORDOR was a cluster-randomized, placebo-controlled trial conducted in Niger’s Dosso region (Figure 1A) to study the effect of twice-annual azithromycin MDA on childhood mortality. The mortality monitoring trial enrolled 594 villages, with 303 assigned to receive azithromycin and 291 assigned to receive a matching placebo twice annually over a 24-month period. A random sample of 30 additional villages from the original sampling frame were enrolled concurrently to monitor the impact of azithromycin MDA on AMR. Villages in the AMR monitoring sample were also randomized to receive azithromycin (N = 15) or placebo (N = 15) (Figure 1B). All children aged 1–59 months residing in trial villages were eligible to receive a dose of azithromycin or placebo twice annually, based on village treatment assignment.

**Figure 1.**
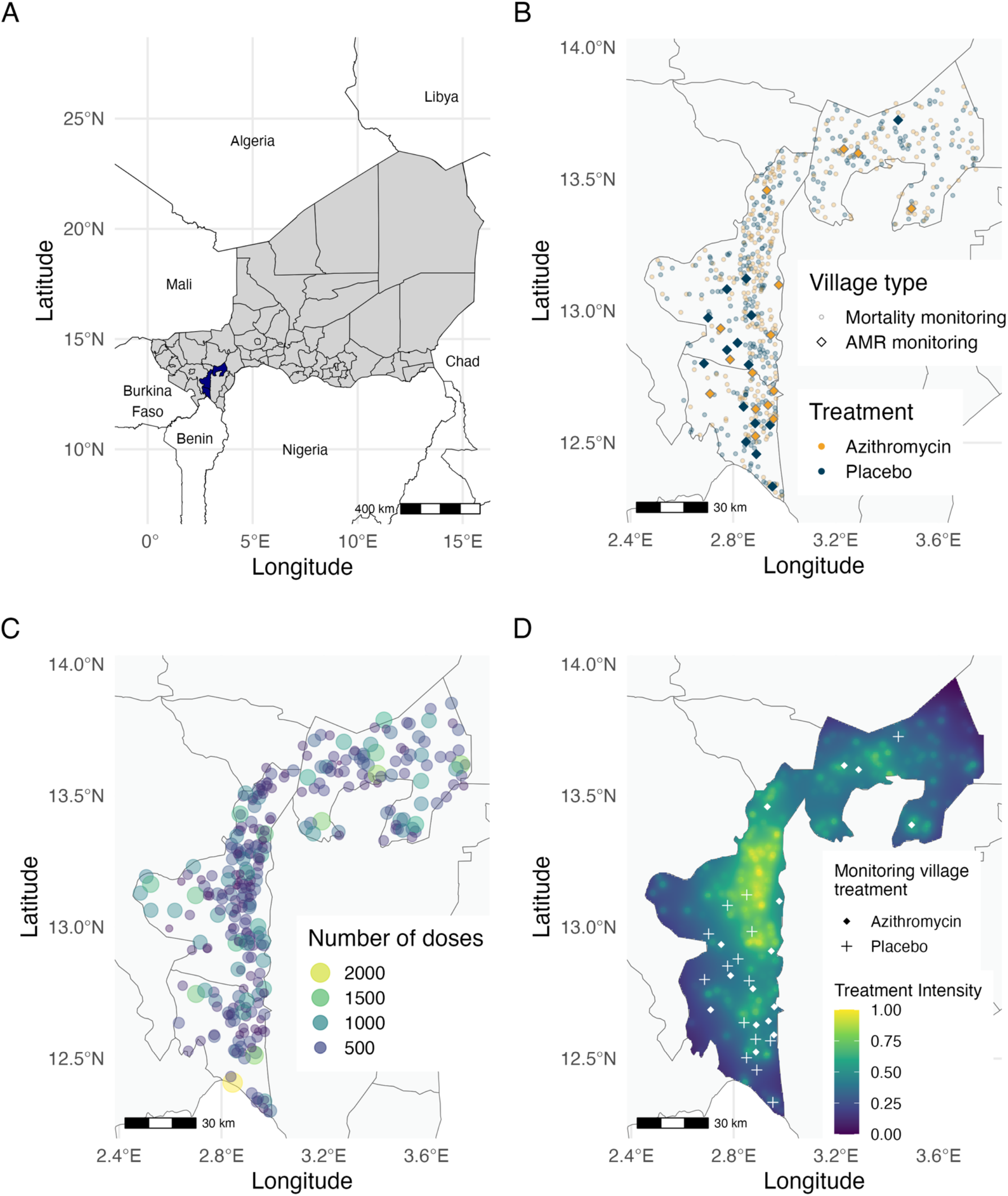
Study area and treatment distribution in the MORDOR trial in Dosso, Niger. A. The study area includes the Boboye, Loga, and Falmey departments (highlighted in blue) in the Dosso region of Niger. The country of Niger is shown in grey, with black lines indicating district boundaries within the grey-shaded area. B. Each point in the study area represents a village enrolled in the trials, with azithromycin-treated villages being represented in yellow and placebo-treated villages in blue. 594 villages (lighter circles) were randomized to receive mass drug administration (MDA) of azithromycin (N = 303) or placebo (N = 291) as part of the MORDOR trial to monitor childhood mortality. 30 other villages (darker diamonds) were separately randomized to receive azithromycin (N = 15) or placebo (N = 15) to monitor anti-microbial resistance. Grey lines depict department boundaries C. Each circle represents a mortality monitoring village treated with azithromycin (yellow circles in panel B). The circle area and color represent the number of doses distributed in the village, with smaller purple circles representing fewer doses and larger yellow circles representing higher number of doses D. The geographic treatment intensity layer was estimated as the inverse distance weighted sum of azithromycin doses distributed in nearby mortality monitoring villages, based on data presented in panel C. Points mark the location of the 30 morbidity monitoring villages, with shapes indicating the treatment assignment. Base maps were sourced from Humanitarian Data Exchange (https://data.humdata.org/) on 9 April 2024. Figure created using script https://osf.io/jrvsw.

We investigated the potential for macrolide resistance to spread beyond treated populations in the context of MDA programs. Mass distribution of azithromycin could exert selective pressure not only within treated populations but also in nearby untreated areas, influenced by patterns of human mobility, environmental factors, and microbial transmission. In the presence of between-village spillover effects, we expect the resistance in a given village to also depend on the number of azithromycin doses distributed in neighboring villages. We hypothesized the effect would be more pronounced in untreated monitoring villages, as these areas would not experience the direct selective pressure of treatment but would still be affected by spillover. We additionally hypothesized that spillover effects could influence AMR in treated monitoring villages by augmenting resistance selection pressure beyond direct treatment. To systematically examine the relationship between spatial variation in azithromycin treatment and resistance selection, we assessed whether AMR increased with higher azithromycin distribution in neighboring villages.

### Geographic variation in azithromycin treatment intensity

We anticipated spatial heterogeneity in the number of antibiotic doses distributed across Dosso due to underlying variation in settlement patterns and the randomized assignment of azithromycin treatment to villages. We estimated an azithromycin geographic treatment intensity variable to represent the potential for geographic spillover of AMR resulting from azithromycin MDA in nearby villages. We quantified geographic treatment intensity at any point in the study region as the cumulative number of doses of azithromycin distributed in mortality-monitoring villages over 24 months (Figure 1C), weighted by the inverse of the distance to that point. We used an inverse distance weighting approach with a linear decay function to balance the influence of nearby and distant treated villages, aligning with the expected antimicrobial resistance spread. There was substantial variation in the geographic treatment intensity variable over the study region and across the 30 monitoring villages (Figure 1D). AMR was measured at baseline (prior to treatment) and again at 24 months, following four rounds of azithromycin distribution, in 30 monitoring villages. Metagenomic deep sequencing was performed on pooled rectal swab samples from 40 children per village, and AMR was quantified as village-level normalized read counts of macrolide– lincosamide–streptogramin (MLS) resistance determinants at AMR-monitoring villages.

### Spillover effects of azithromycin MDA to nearby villages

MLS resistance determinants remained low and at baseline levels in placebo-treated AMR monitoring villages at 24 months, regardless of azithromycin geographic treatment intensity (Figure 2A, 2B). We quantified the association between MLS resistance and azithromycin geographic treatment intensity using Spearman rank-order correlation. To assess statistical significance, we applied permutation tests in which we permuted the treatment assignments of surrounding mortality-monitoring villages, using a conditional permutation approach [20,21]. Azithromycin treatment intensity was uncorrelated with macrolide resistance at both timepoints (P = 0.99 at baseline, P = 0.83 at 24 months) in placebo-treated villages. These results are consistent with no geographic spillover of azithromycin-induced resistance in untreated monitoring villages.

**Figure 2.**
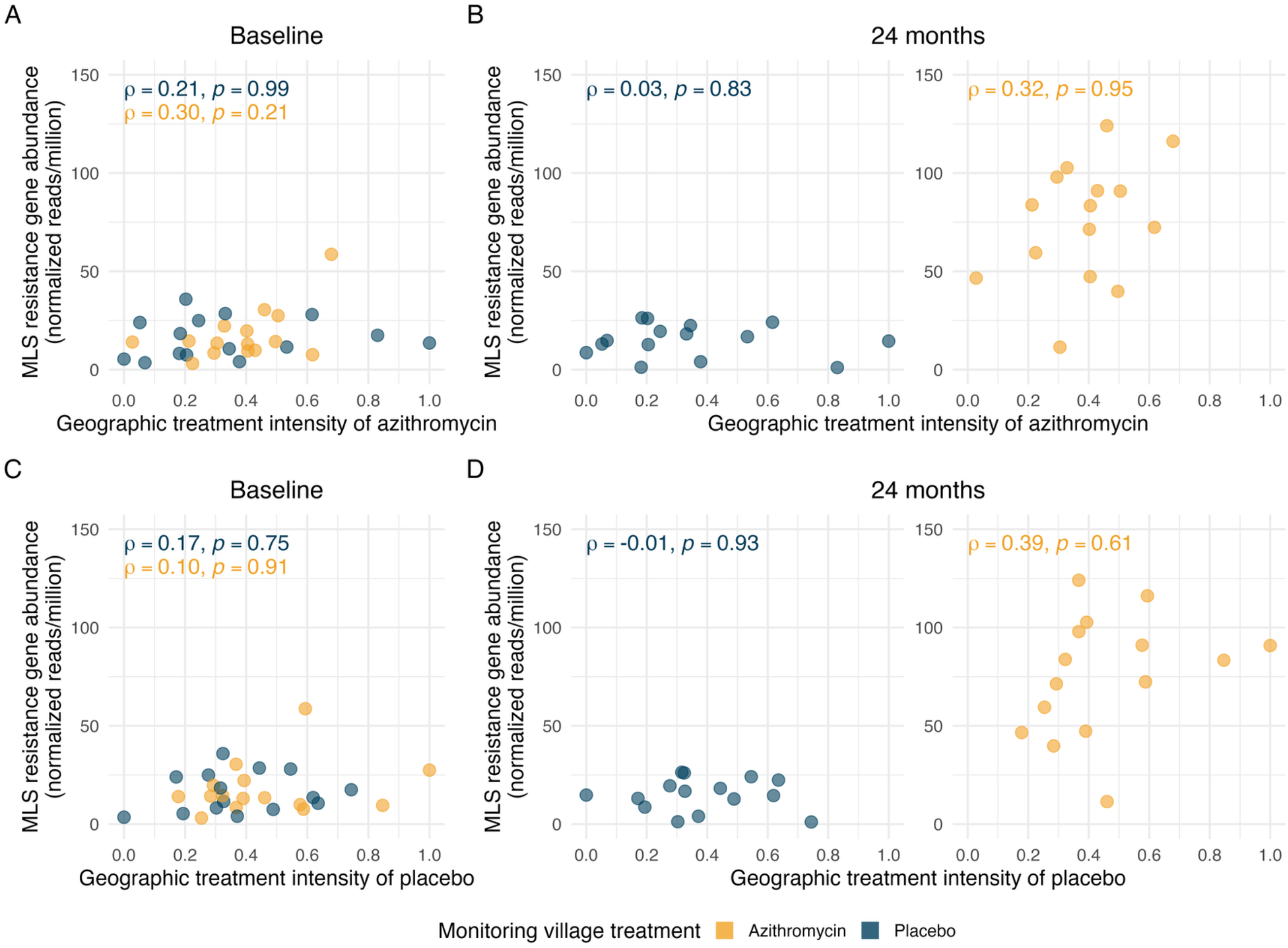
Spillover effects of mass drug administration (MDA) with azithromycin on genotypic resistance to Macrolides–lincosamides–streptogramins (MLS) A. Relationship between the azithromycin treatment intensity and the village-level normalized reads of MLS resistance determinants (in reads per million) at baseline, estimated as Spearman rank-order correlations (ρ). Points represent anti-microbial resistance (AMR) monitoring villages, with azithromycin-treated villages in yellow and placebo-treated villages in blue. B. Relationship between the azithromycin treatment intensity and the village-level normalized reads of MLS resistance determinants (in reads per million) at 24 months with placebo-treated villages in the left panel and azithromycin-treated villages on the right. Left panel: In placebo-treated villages, MLS AMR levels remained low across all values of geographic treatment intensity, indicating no evidence of spillover of resistance following MDA. Right panel: Higher MLS AMR levels were observed in the azithromycin-treated villages at 24 months as compared to the baseline. Spearman correlations in azithromycin-treated villages remained similar at baseline (ρ = 0.30) and at 24 months (ρ = 0.32), indicating a stable relationship across time points. C. Relationship between the placebo treatment intensity (negative control exposure) and the village-level normalized reads of MLS resistance determinants at baseline. D. Relationship between the placebo treatment intensity (negative control exposure) and the village-level normalized reads of MLS resistance determinants at 24 months with placebo-treated villages in the left panel and azithromycin-treated villages on the right. Left panel: In placebo-treated villages, MLS AMR levels remained low across all values of placebo treatment intensity, with no significant correlation being observed. Right panel: Similar to panel B, Spearman correlations in azithromycin-treated villages indicate a monotonic trend of MLS AMR with placebo treatment intensity. The effect of placebo treatment intensity being similar to azithromycin treatment intensity indicates that the observed modest and non-significant correlation in panel B is likely not attributable to azithromycin distributions. All p-values are two-sided and derived from a permutation test. Figure created using script https://osf.io/4puya.

As reported previously [4], azithromycin-treated AMR-monitoring villages experienced a sharp increase in MLS resistance determinants from baseline to 24 months (Figure 2A, 2B), attributable to the direct effects of azithromycin MDA. A modest correlation between geographic treatment intensity and MLS resistance in azithromycin-treated villages was observed at baseline (ρ = 0.30) and remained similar at 24 months (ρ = 0.32), though neither was significant under a conditional permutation test (P = 0.21 at baseline, P = 0.95 at 24 months). It is therefore likely that the observed, modest correlation reflects pre-existing spatial patterns rather than an emergent spillover effect of azithromycin MDA.

### Negative control exposure analysis

As an additional robustness check, we estimated the geographic treatment intensity of placebo doses as a negative control exposure (Figure 2C, 2D). Using the same methods used to estimate azithromycin geographic treatment intensity, but with the cumulative number of placebo doses distributed instead of the cumulative azithromycin doses, the negative control exposure analysis was designed to detect spurious associations arising from unmeasured confounding, or other non-causal sources of variation in AMR that are unrelated to azithromycin MDA [22,23]. As with the geographic treatment intensity of azithromycin, we observed a modest positive correlation between MLS resistance determinants and geographic treatment intensity of placebo in azithromycin-treated monitoring villages at 24 months (ρ = 0.39); however, this association was also not statistically significant under a conditional permutation test (*P =* 0.61, Figure 2D). Since placebo treatment is not expected to have any biological impact on MLS resistance determinants, the similarity of the negative control analysis and the primary analysis suggests that the observed non-significant correlations are unlikely to result from the spillover effects of azithromycin MDA. We estimated a partial Spearman rank correlation between geographic treatment intensity of azithromycin and AMR, conditioning on the negative control exposure (partial ρ = 0.1, P = 0.7) in azithromycin-treated AMR-monitoring villages at 24 months. The unadjusted correlation was already weak and non-significant (ρ = 0.32, P = 0.99) and further decreased in magnitude after conditioning on the negative control, reinforcing that geographic variation in azithromycin MDA intensity is unlikely to be a driver of AMR patterns.

### Non-parametric analysis of geographic spillovers

To further investigate spatial spillover by isolating the effect of distance from azithromycin-treated mortality monitoring villages, we estimated the association between azithromycin exposure and MLS resistance within discrete distance bands. Specifically, we calculated the cumulative number of azithromycin doses distributed within 10 km, 10–20 km, and 20– 30 km of each AMR monitoring community over 24 months (Figure 3A, 3B) and assessed their association with MLS resistance using Spearman rank-order correlation; statistical significance was evaluated using a permutation test. In the presence of AMR spillover, we hypothesized that there would be a stronger correlation between azithromycin doses delivered and MLS resistance at smaller distances since closer proximity increases the likelihood of transmission of resistant strains between villages.

**Figure 3.**
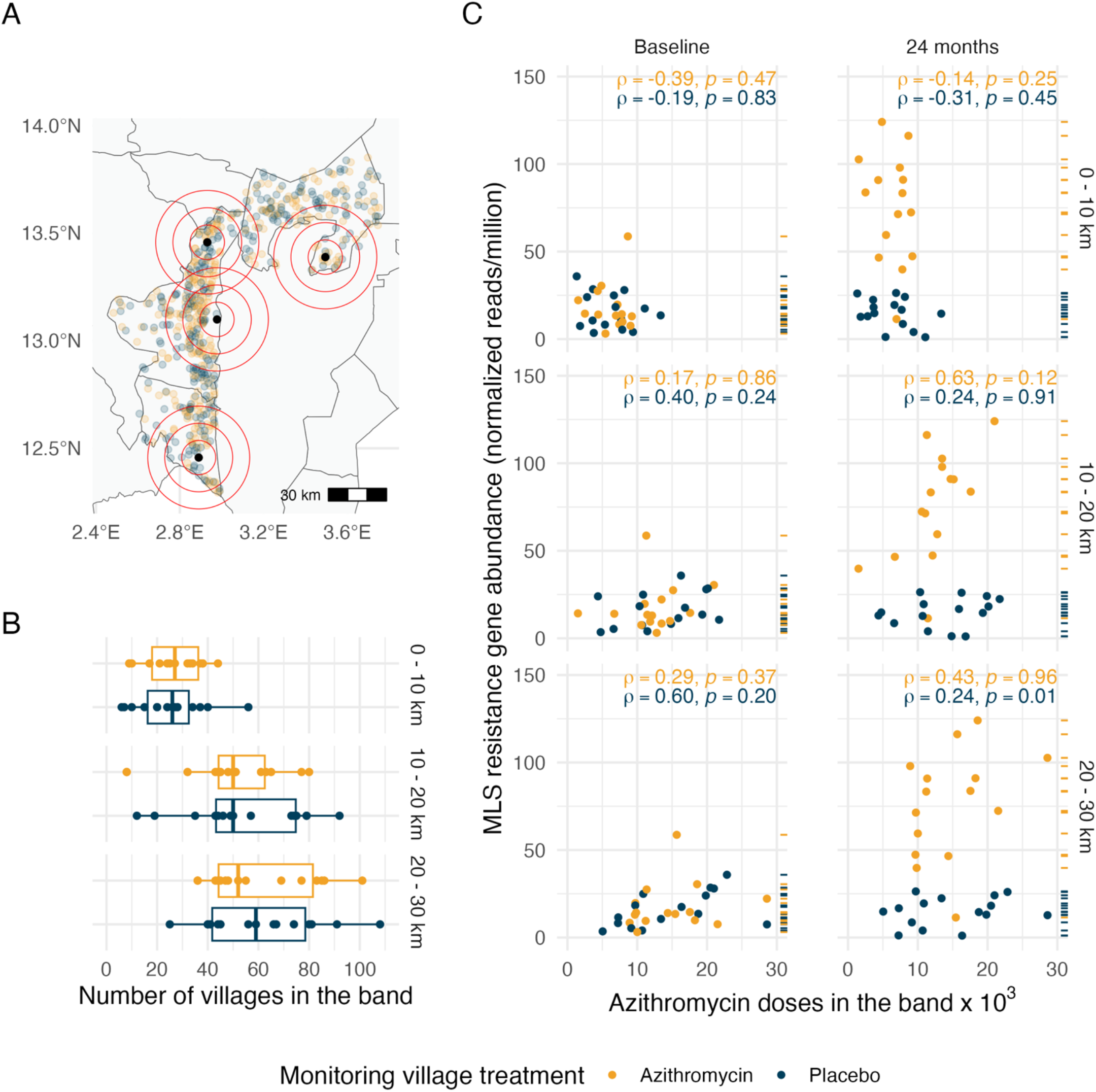
Non-parametric assessment of spillover effects of azithromycin MDA on genotypic resistance to Macrolides–lincosamides–streptogramins (MLS) by 10km geographic bands. A. Map of the study area indicating the locations of 4 of 30 AMR monitoring villages (black circles) for illustration and all 594 mortality monitoring villages, with azithromycin-treated villages in yellow and placebo-treated villages in blue. Red concentric circles represent distance bands at 0–10 km, 10–20 km, and 20–30 km centered at AMR monitoring villages, used to assess spillover effects of azithromycin treatment intensity. B. Number of mortality monitoring villages within each distance band from the AMR monitoring villages. Boxplots show the distribution of villages within each band, stratified by treatment assignment of the AMR monitoring villages. As expected from randomization, the distributions are similar for azithromycin-treated and placebo-treated AMR monitoring villages. C. Association between azithromycin doses administered within each distance band (in thousands) and the normalized abundance of MLS resistance determinants (reads per million) in AMR monitoring villages at baseline (left) and 24 months (right). Each row of scatter plots corresponds to a specific distance band: 0–10 km (top), 10–20 km (middle), and 20–30 km (bottom). Dashes on the right of each panel indicate the normalized abundance of MLS resistance determinants of each village in the panel. Spearman rank-order correlations (ρ) with p-values from permutation tests are shown for both time points. The absence of stronger correlations at closer distances at 24 months suggests no evidence of spillover effects on MLS resistance. Figure created using script https://osf.io/ka68w.

The correlations between MLS resistance determinants and geographic treatment intensity within most distance bands were low. These results also suggest no evidence of a significant increase in MLS resistance associated with a higher number of doses distributed in proximate villages. Although there was a significant correlation in placebo-treated villages, in the 20-30 km band, this was not accompanied by distance-based trends that would suggest geographical spillover of AMR to nearby villages (Figure 3C).

### Sensitivity analyses

Gravity models, commonly used to describe human movement and infectious disease dynamics [24], were adapted to assess macrolide resistance spread by using an inverse distance-squared weighted sum of azithromycin doses in mortality monitoring villages as an alternative geographic treatment intensity measure. The results of this sensitivity analysis were similar to the primary analysis (Supplementary Figure 1), indicating the robustness of the primary analysis to the azithromycin treatment intensity measure used. Additionally, the results were also robust to a leave-one-out analysis, verifying that the findings were not driven by individual AMR monitoring villages (Supplementary Figure 2).

The pre-specified analysis based on a permutation test could not detect a significant effect of MDA on spillover of macrolide resistance. We conducted an additional, post hoc analysis to estimate the fold-change in resistance genes corresponding with an increase of azithromycin doses in a 10 km radius surrounding a village. Unlike the conditional permutation test, inference no longer relies exclusively on the randomization, but it provides a complementary assessment of the precision around the observed null effect. In log-linear models adjusted for baseline macrolide resistance load, as well as for azithromycin doses distributed within the 10–20 km and 20–30 km distance bands, we estimated that an increase of 5,000 azithromycin doses within a 10 km radius (corresponding to the median value of observed doses) would result in a fold change in macrolide resistance genes of 0.7 (95% CI: 0.2 to 1.9) in placebo-treated villages and 1.0 (95% CI: 0.4 to 2.3) in azithromycin-treated villages. Point estimates are consistent with the randomization-based inference (no spillover) and the upper limits of the intervals show that modest increases in resistance genes, up to a 2.3-fold increase, are unlikely but compatible with the data given the relatively small number of 15 monitoring villages in each analysis.

We additionally assessed the relationship between the measure of geographic treatment intensity and independent measures of population density and distance to primary health post to help provide additional context for the results. High-resolution population estimates [25] were associated with geographic treatment intensity and MLS resistance (Supplementary Fig 4), illustrating that observational analyses that fail to account for population density could be confounded. Re-fitting the log-linear models to estimate fold-changes, adjusting for population density in addition to baseline macrolide resistance load and azithromycin doses within the 12–20 km and 20–30 km distance bands, led to estimates of 0.3 (95% CI 0.1 to 1.3) in placebo-treated villages and 0.7 (95% CI 0.2 to 1.9) in azithromycin treated villages. Proximity to the nearest health center [26], which could influence access to antibiotics and selection pressure, was not associated with either geographic treatment intensity or AMR prevalence (Supplementary Fig 5).

Finally, we extended our analysis to investigate potential spillover effects on genotypic resistance to other classes of antibiotics beyond MLS. No significant relationship was observed between geographic treatment intensity and non-macrolide antimicrobial resistance determinants at either baseline or 24 months (Supplementary Figure 6). These findings align with previously reported results, which found no significant between-group differences in resistance for non-macrolide antimicrobial classes at 24 months [4].

## Discussion

This study found no evidence of between-village geographical spillover of macrolide AMR to nearby untreated villages following large-scale, twice-annual azithromycin MDA to children aged 1-59 months old in Niger. Genetic load of MLS resistance in placebo-treated monitoring villages remained at baseline levels after 24 months of intervention and had no association with the number of azithromycin doses administered in nearby treated villages.

These results do not support the hypothesis that azithromycin MDA-induced selection of AMR extends beyond treated villages at the geographic scales evaluated. Substantial, unmeasured indirect effects of mass drug administration on resistance are improbable beyond the direct selection for MLS resistance observed within treated communities over a 24-month period (four MDA treatments). Our findings suggest that current AMR monitoring strategies in sentinel villages, which do not explicitly account for spillover effects, are likely sufficient to capture the extent of AMR selection attributable to MDA during the initial years of program implementation. Internally consistent results across multiple analytical methods reinforce our inference and demonstrate that effects on AMR selection estimated in the original trial analysis are unlikely to be biased by between-cluster spillover. This analysis provides new insights into the spatial scale of azithromycin treatment variation and AMR selection following MDA and contributes to evidence-based decision-making regarding the use of azithromycin MDA for reducing childhood mortality in high-risk settings.

Unlike conventional randomized trials where outcomes are measured among treated and untreated units, in this analysis the MDA treatments used to quantify spillover effects were delivered to different clusters (594 villages) than those used to measure AMR outcomes (30 monitoring villages), a generalization of the randomized controlled trial [28]. Cluster randomization over 594 villages created variation in geographic treatment intensity across the random subsample of 30 AMR monitoring villages, and permuting the random assignments using a conditional permutation test allows for an exact, non-parametric test for the presence of spillover effects between villages [20,21]. Along with failure to reject the null under the conditional permutation test, the separate finding of a similar correlation between geographic treatment intensity and AMR, regardless of whether geographic treatment intensity was estimated using azithromycin or placebo doses (Figure 2B, 2D), further supports the view that the observed correlation is due to factors other than spillover. Robustness checks that adjusted for local population density and distance to health center did not change our inference and reinforced the conclusion that geographic treatment intensity did not independently influence AMR in this study.

The analysis has several caveats. Monitoring village AMR measurements relied on a random sample of 40 children per village, providing a good measure of AMR genetic load in each location but the relatively small number of AMR monitoring villages (N = 30) may limit our ability to fully capture the range of azithromycin treatment intensity across the study area. Future studies that include a larger set of monitoring communities could have higher power to detect spillover effects, if present. Another potential limitation is the choice of spillover measure: we assume the potential for AMR spillover is proportional to the absolute number of azithromycin doses distributed rather than the proportion of the population treated. The distance decay is also assumed to be uniform across the study area, without accounting for potential variations in ease of connectivity and travel time between villages or geographic factors such as terrain and remoteness. This assumption may oversimplify some spatial relationships. For this analysis, we focused on genotypic resistance, the presence of resistance genes, rather than further downstream outcomes such as phenotypic resistance or clinical impact. While genotypic resistance may not directly indicate treatment failure, it serves as an early indicator of resistance potential within the microbial population. Thus, if we do not observe a discernible spillover effect in genotypic resistance, it is unlikely that we will find one further downstream. Finally, this study provides insights into local geographic spillover effects over a 24-month period, but azithromycin MDA has continued in the Dosso region for several years beyond this study [29], and the use of azithromycin MDA in trachoma control programs regularly spans several years. Longer-term monitoring of AMR trends in regions with repeated rounds of MDA would help measure cumulative effects on resistance over time, including potential geographic spillover effects. Future analyses of large-scale, azithromycin MDA trials in the region [2,3,30] could enable additional assessments over longer periods.

## Conclusion

In summary, there was no evidence of between-village spillover of macrolide resistance following azithromycin MDA, suggesting that AMR effects are largely localized to treated villages without spreading to neighboring areas. This study highlights the value of randomized trial designs to test for spillover effects of large-scale public health interventions even if outcome monitoring is focused in a subsample of units. Future studies should continue to explore the longer-term impacts of repeated azithromycin MDA on selection and possible geographic spillovers of AMR.

## Methods

This is a secondary analysis based on the MORDOR mortality trial (NCT02047981), a cluster-randomized trial that evaluated the impact of twice-yearly mass distribution of oral administration of azithromycin on childhood mortality, and its sister trial, MORDOR Morbidity trial (NCT02048007). The geographic spillover analysis plan was pre-specified (https://osf.io/xyqnr).

### Trial oversight

The trial protocol was reviewed and approved by the Committee for Human Research at the University of California, San Francisco (protocol #10-01036) and the Ethical Committee of the Niger Ministry of Public Health. The trial was monitored by an independent Data and Safety Monitoring Committee and was conducted in accordance with the Declaration of Helsinki. Due to low literacy in Niger, trial oversight committees approved the use of oral consent from guardians of children before administration of treatment and each sample collection.

### Study population

The trial was conducted in the Boboye, Loga and Falmey departments in Dosso region of Niger (Figure 1A). All non-urban villages with an estimated population of 200-2000 people in the study area were eligible to be included in the trial. Eligible villages were randomized to receive MDA with azithromycin or placebo to monitor the impact of azithromycin MDA on childhood mortality. Additionally, a random sample of 30 other villages from the same pool of eligible villages were separately randomized to receive MDA of azithromycin or placebo to monitor the impact of MDA on AMR (Figure 1B). All children aged 1–59 months in enrolled villages received the assigned treatment, azithromycin or matching placebo, twice annually for 2 years. Baseline AMR measurements were collected in the AMR-monitoring villages prior to the first MDA distribution, starting in December 2014, and follow-up AMR measurements were collected at 24 months, six months after the fourth MDA distribution.

### Geographic Treatment Intensity

In this context, geographic treatment intensity represents the potential for geographic between-village spillover of AMR resulting from azithromycin treatment in nearby clusters. A geographic treatment intensity variable was computed based on the cumulative number of doses of azithromycin distributed as part of the mortality monitoring trial. Geographic treatment intensity was estimated as an inverse distance weighted sum of the number of doses of azithromycin distributed at each AMR-monitoring location to allow for higher weights for nearby treated villages while accounting for diminishing influence with increasing distance. If the number of azithromycin doses distributed at locations x_1_,…,x_n_ are v_1_,…,v_n_ respectively, then the geographic treatment intensity at a location u is g(u)= ∑w_i_v_i_ where the weights are the inverse p^th^ powers of distance, w_i_=1/d(u,x_i_)^p^ where d(u,x_i_)=∣∣u−x_i_∣∣ is the Great circle distance from u to x_i_. As the power is reduced, the smoothness of the interpolated spatial distribution increases.

The primary analysis was done using a power of 1, and a sensitivity analysis was done using a power of 2. A power of 1 in the inverse distance weighting calculation was selected for the primary analysis as it provides a balance between capturing the influence of nearby treated villages while maintaining contributions from more distant villages, analogous to Shepard’s method [31] but applied to weighted sums rather than interpolated averages. This linear decay function maintains relatively high influence within the first several kilometers before diminishing at greater distances, which aligns with the expected pattern of antimicrobial resistance dissemination across geographic areas [32]. Gravity models are widely used to describe human movement across spatial scales and have recently been applied to model population mixing and mobility in infectious disease transmission dynamics [24,33]. We adapted the principles of gravity models to evaluate the spatial dissemination of macrolide resistance by assessing the inverse distance-squared weighted sum of the cumulative number of azithromycin doses distributed in mortality monitoring villages as a sensitivity analysis.

In instances when treated village distances fell below 1 km from monitoring villages, distances were truncated at 1 km to avoid inordinately large weights in the calculation. The geographic treatment intensity variable was rescaled to range from 0 to 1 by dividing by its maximum value. AMR monitoring villages were assigned the geographic treatment intensity value at their geographic centroid.

In this geographical treatment intensity calculation, we assume that the potential for AMR spillover is proportional to the absolute number of azithromycin doses distributed rather than the proportion of the population treated. Additionally, this approach does not account for the possibility that larger AMR monitoring villages might experience more spillover events due to increased human mobility and bacterial transmission or, conversely, that smaller AMR monitoring villages might exhibit more measurable spillover effects due to lower background resistance levels. However, there was no clear association between AMR-monitoring village size and either geographic treatment intensity (Supplementary Figure 3) or MLS resistance gene abundance (Supplementary Figure 7), suggesting that village size did not systematically influence the observed outcomes. Furthermore, we assume that geographic distance alone dictates the strength of spillover effects, without incorporating other potential modifying factors such as population density, healthcare access, or environmental pathways of resistance transmission.

### Village load of antimicrobial resistance genes

Rectal swabs were collected from a random sample of 40 children aged 1 to 59 months in each AMR monitoring village at baseline and at 24 months. Samples from each village, at each time point, were pooled. DNA was extracted from the pooled sample, and its concentration was quantified and normalized for sequencing library preparation. Libraries were sequenced with 150-nucleotide paired-end reads. Reads underwent three rounds of human sequence removal, quality filtering, and taxonomic classification before nonhuman reads were aligned to the MEGARes database to identify antibiotic-resistance determinants with a gene fraction >80%. Microbiome sequencing reads can be accessed at https://microbiomedb.org/mbio/app. The normalized abundance of these determinants (measured as reads per million) was calculated for each pooled sample [4,5]. Community-level averages of MLS resistance determinants were used as the primary outcome measure.

Similarly, normalized abundance of resistance determinants was estimated for other classes of antibiotics, namely, aminocoumarins, aminoglycosides, bacitracin, beta-lactams, elfamycins, fluoroquinolones, fosfomycin, glycopeptides, metronidazole, phenicol, rifampin, sulfonamides, tetracyclines, trimethoprim, and multidrug resistance.

### Spillover effect assessment

To assess the potential spillover effects of azithromycin MDA on MLS resistance, the association between azithromycin treatment intensity and the normalized abundance of MLS resistance determinants was evaluated. The primary analysis focused on placebo-treated monitoring villages, testing the hypothesis that resistance in these villages was associated with the geographic treatment intensity of azithromycin in surrounding villages. Additionally, the association within azithromycin-treated monitoring villages was examined to explore whether a similar relationship existed despite the direct effects of treatment.

Spearman rank-order correlations were used to evaluate these associations, stratified by whether the monitoring villages received placebo or azithromycin. Analyses were conducted at baseline and 24 months post-MDA to account for temporal dynamics. To ensure the robustness of the correlations estimated, a leave-one-out analysis was conducted, sequentially excluding each AMR monitoring village from the dataset to evaluate the influence of individual villages on the overall results. In addition to MLS resistance, we also assessed resistance determinants for other antibiotic classes to detect broader spillover effects.

To test the statistical significance of observed correlations between geographic treatment intensity and MLS resistance, we used a non-parametric, conditional permutation test to detect the presence of spillover effects between villages [20,21]. In each permutation, the treatment labels of the 594 mortality monitoring villages were re-randomized, and the geographic treatment intensity variable was re-estimated for the AMR monitoring villages. AMR monitoring village treatments were held fixed in the permutation tests, creating a distribution for the correlation between the monitoring village AMR load and surrounding geographic treatment intensity under the null hypothesis of no between-village spillover effects [20,21]. The null distribution for all test statistics was generated from 1000 permutations. Observed correlations were compared to the null distribution to determine two-sided p-values, with significance assessed at an alpha of 0.05.

### Negative control exposure analysis

The placebo treatment in mortality-monitoring villages served as a negative control exposure to assess robustness. We repeated the geographic spillover analysis, substituting the cumulative number of placebo doses for azithromycin doses when estimating the geographic treatment intensity, using the same inverse distance-weighted approach and identical testing procedures. Any observed associations between the geographic treatment intensity of placebo and AMR load could be attributed to confounders unrelated to azithromycin exposure.

### Non-parametric treatment measure

To separate the effect of distance and the number of doses, a non-parametric distance-based analysis was conducted. Considering each AMR monitoring community as the center, concentric non-overlapping distance bands set at 0–10 km, 10–20 km, and 20–30 km were established. The cumulative number of azithromycin doses administered within each band was used as an alternative azithromycin treatment intensity estimate within each ring. The number of azithromycin doses within each distance band was examined as a predictor of MLS resistance determinants in the AMR monitoring community at baseline and 24 months. Associations between azithromycin doses in each distance band and MLS resistance determinants were estimated using Spearman rank-order correlations and their significance was evaluated using a permutation test.

### Additional sensitivity analyses

The pre-specified analysis focused on the randomization in the design and conditional permutation test for inference. After review of the main results and negative control analysis, we conducted three additional model-based sensitivity analyses as robustness checks (not pre-specified). First, we fitted log-linear models relating normalized macrolide resistance gene load to the number of azithromycin doses within 0–10 km, 10–20 km, and 20–30 km of each village, adjusting for baseline macrolide resistance levels. From these models, we estimated the fold-change in macrolide resistance gene abundance associated with an increase of 5,000 doses within 10 km. We fit separate models for placebo-treated and azithromycin-treated monitoring villages, and estimated model-based 95% confidence intervals and P-values for the fold-change. Second, we evaluated the association between two geographic characteristics that could be correlated with our measure of geographic treatment intensity and load of antimicrobial resistance: local population density and distance to the nearest health center. We characterized local population density using data from the Meta High Resolution Settlement Layer [25] and summarized the number of children under 5 years within the distance rings of 0-10 km, 10-20 km, and 20-30 km of each AMR-monitoring village. We also estimated distance to the nearest Centres de Santé Integré facility, a primary health center that is equipped to provide basic inpatient and outpatient care. After determining that population density was associated with both geographic treatment intensity and macrolide resistance, we included it as an additional covariate in the log-linear model.

All analyses were performed using R version 4.4.0.

## Data Availability

Data and replication files are available through the Open Science Framework (https://osf.io/qxtec/). Village geographic coordinates have been omitted to protect participant confidentiality.

## Acknowledgments

The MORDOR trial was supported by a grant from the Gates Foundation (OP1032340 to TML). This work was additionally supported in part by the National Institute of Allergy and Infectious Diseases (R01AI175250 to KO and TML, R01AI158884 to BFA, R01AI166671 to BFA, R35GM147702 to SB) and the Centers for Disease Control and Prevention (CDC U01CK000590 to AS and SB, as part of the Modeling Infectious Diseases in Healthcare Network). Pfizer provided both the azithromycin and the placebo oral suspensions.

