## Supplementary Information File for "Geographic spillover of antimicrobial resistance from mass distribution of azithromycin"

Ariktha Srivathsan et al.

### Supplementary Information File

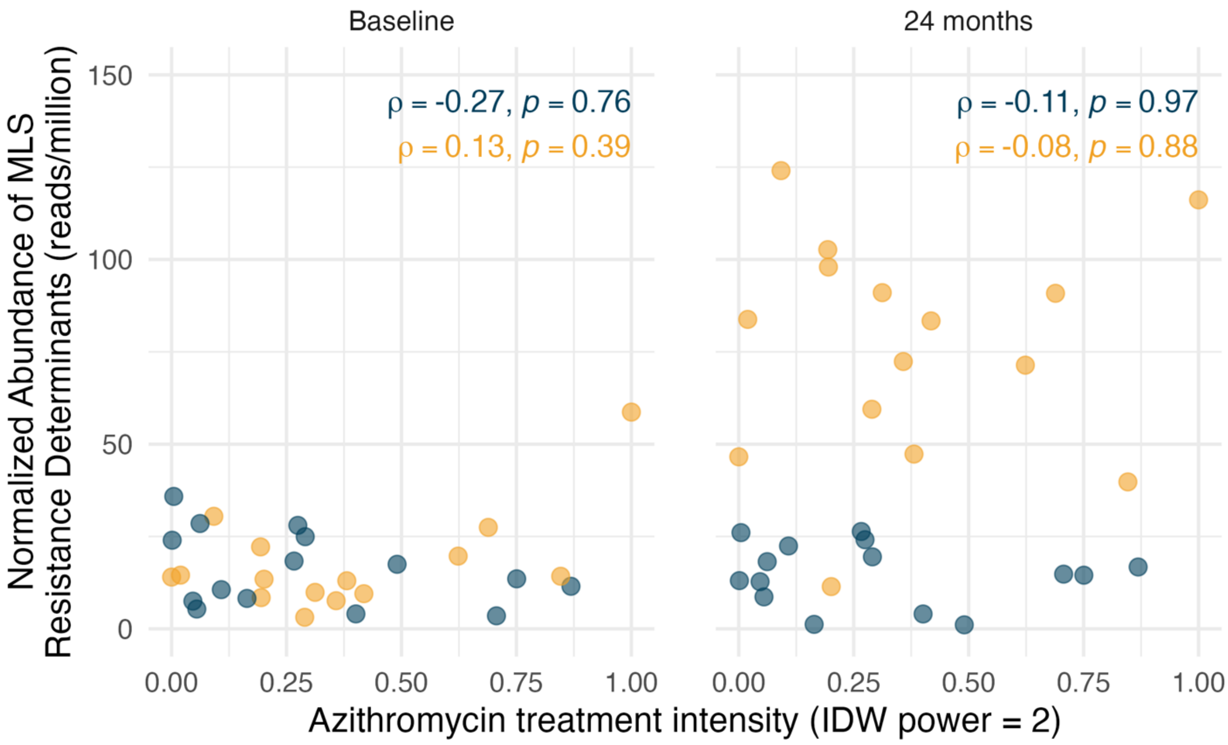


**Supplementary Figure 1: Sensitivity analysis of the relationship between azithromycin treatment intensity and the abundance of Macrolides–lincosamides–streptogramins (MLS) resistance determinants, using an alternative measure of geographic treatment intensity.**

This analysis adapts gravity models, widely used to describe human movement and spatial dynamics, to quantify geographic treatment intensity. The measure was calculated as the inverse distance-squared weighted sum of cumulative azithromycin doses distributed in mortality monitoring villages, providing a metric to evaluate the spatial dissemination of macrolide resistance.

Scatterplots show the normalized abundance of MLS resistance determinants (reads per million) in 30 AMR monitoring villages. Results are presented at baseline (left column) and 24 months (right column) and are stratified by treatment arm: azithromycin-treated villages (yellow points) and placebo-treated villages (blue points). Results align with the primary analysis, showing no systematic associations between azithromycin treatment intensity and resistance determinants at 24 months. These findings reinforce the robustness of the primary analysis, highlighting the localized nature of azithromycin-mediated resistance selection and minimal spillover effects, regardless of the geographic treatment intensity measure used. Figure created using script https://osf.io/v4p5e.

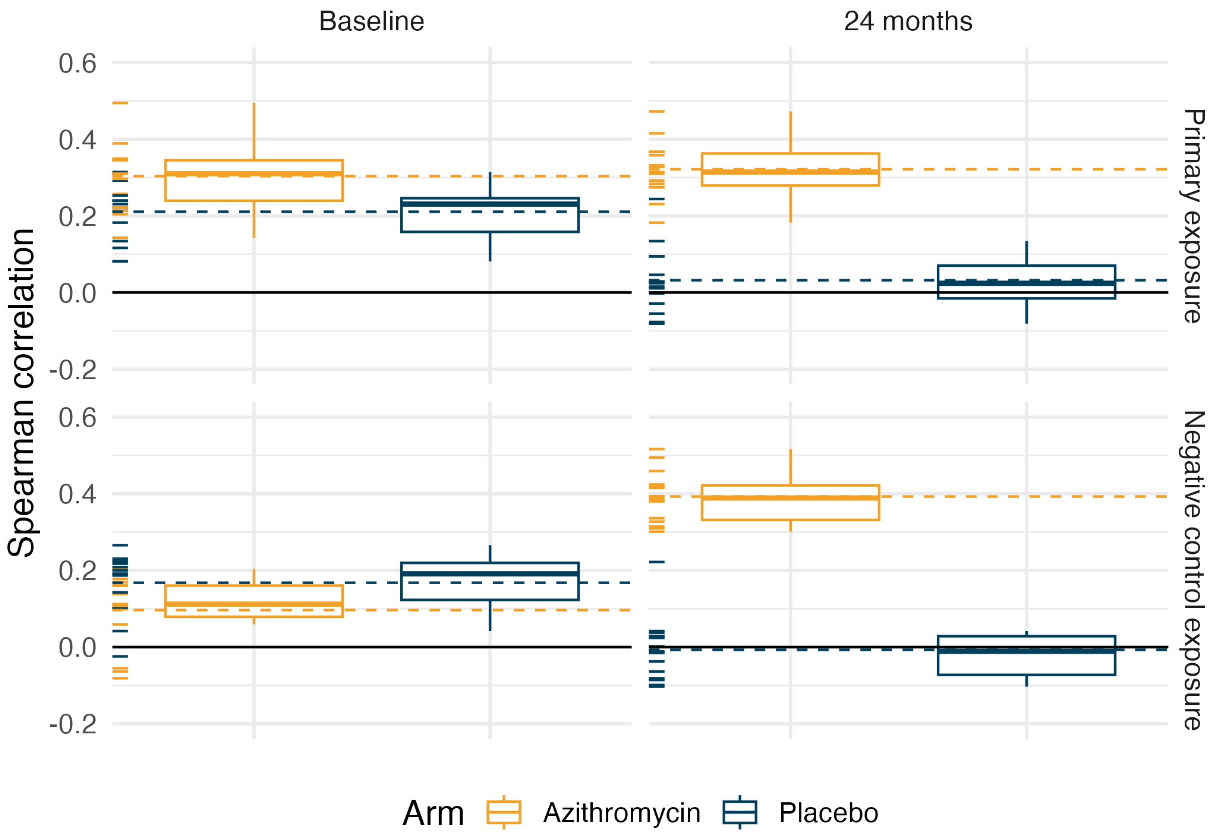


**Supplementary Figure 2: Leave-one-out analysis of the Spearman correlation between azithromycin doses and MLS resistance determinants across study arms and time points.**

Each AMR monitoring village was sequentially removed from the dataset to evaluate the influence of individual villages on the overall results. Boxplots depict the distribution of Spearman correlations, stratified by treatment arm (azithromycin vs. placebo) and time point (baseline vs. 24 months) to evaluate consistency within each group. Dashed lines represent the observed correlation for each group, while the dashes on the left of each panel indicate the observed Spearman correlation for each subset with one village excluded.

Top row (Primary result): Correlations for the relationship between azithromycin treatment intensity and normalized MLS resistance gene abundance.
Bottom row (Negative control): Correlations for the relationship between placebo treatment intensity and normalized MLS resistance gene abundance.

The distributions of correlations were not substantially influenced by the exclusion of any individual village, underscoring the robustness of the analysis. Figure created using script https://osf.io/v4p5e.

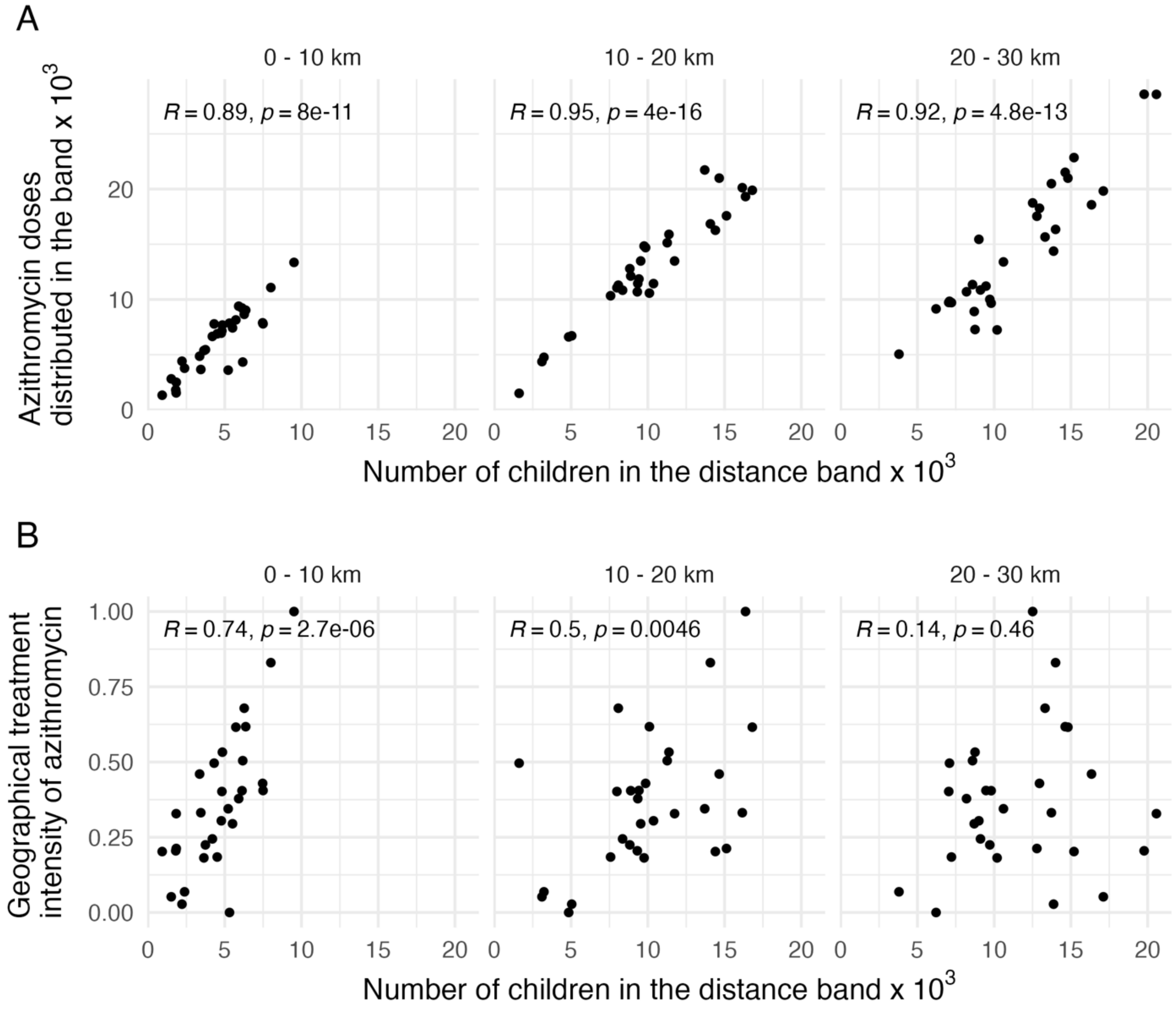


**Supplementary Figure 3: Relationship between the number of children in distance bands and azithromycin treatment metrics at the 30 AMR monitoring villages.**

A. Correlation between the number of children in the 0–10 km, 10–20 km, and 20–30 km distance bands and the total number of azithromycin doses distributed in those bands. Strong positive correlations are observed in all distance bands, with high Pearson correlation values (R).

B. Correlation between the number of children in each band and the geographical treatment intensity of azithromycin, calculated as the inverse-distance weighted sum of the doses of azithromycin distributed in mortality monitoring communities. As expected from the inverse-distance weighting approach, the correlation weakens progressively with increasing distance from the AMR monitoring village. Moderate correlations are observed for the 0–10 km and 10–20 km bands, while the correlation weakens substantially in the 20–30 km band. Figure created using script https://osf.io/v4p5e.

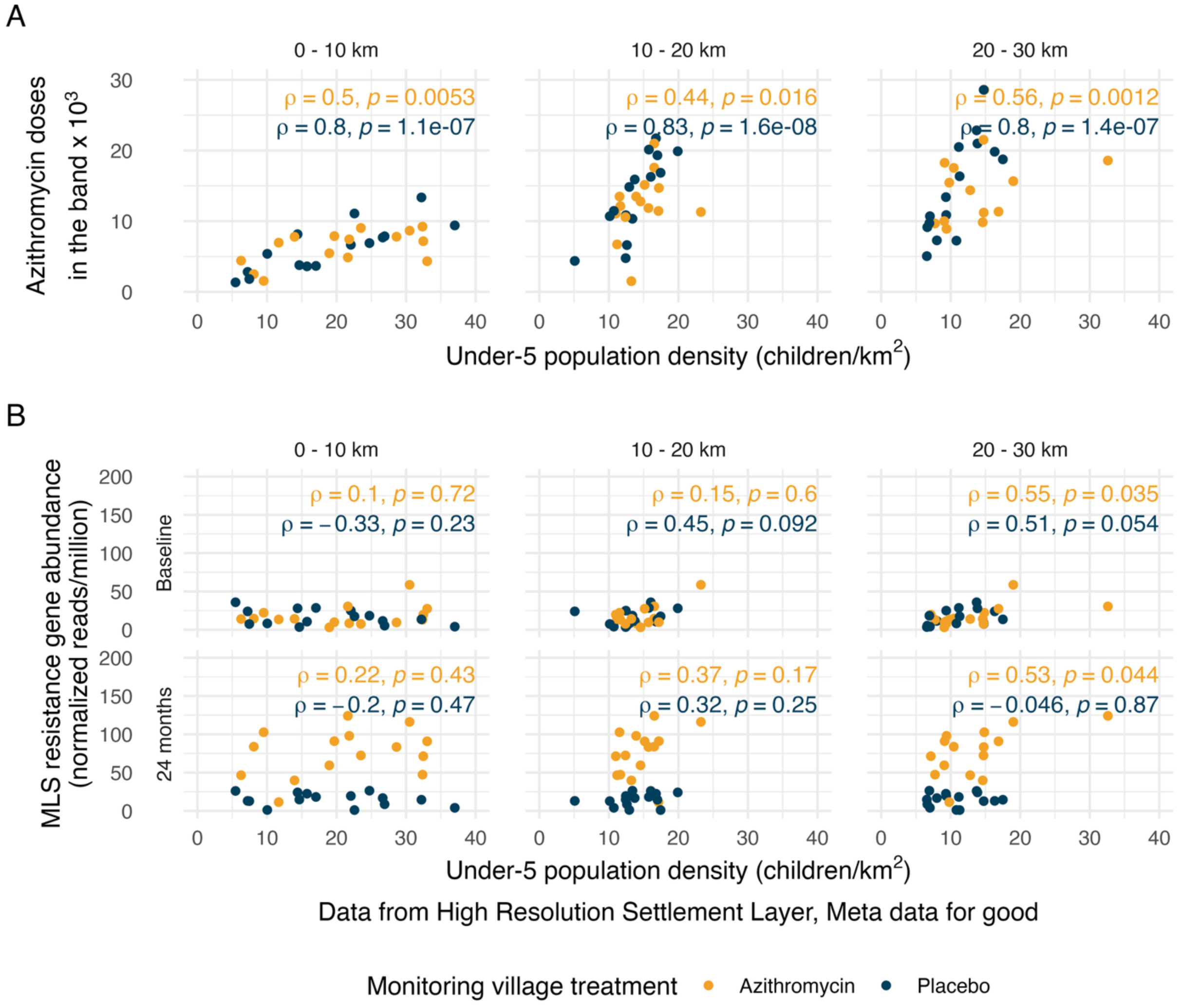


**Supplementary Figure 4: Relationship between azithromycin MDA and subsequent MLS resistance by population density.**

(A) Spearman correlation between under-five population density and the total number of azithromycin doses distributed within 0–10 km, 10–20 km, and 20–30 km distance bands surrounding each AMR monitoring village. Strong positive statistically significant correlations are observed between population density and azithromycin doses distributed in each band. This finding aligns with expectations, as under high MDA coverage, the number of doses distributed is inherently linked to the number of children in the area, which, in a fixed geographic region, corresponds directly to population density.

(B) Relationship between under-five population density and MLS resistance in AMR monitoring villages at baseline (top row) and 24 months (bottom row). At baseline and 24 months, statistically significant Spearman's correlations are observed in the 20-30 km distance band in azithromycin-treated AMR monitoring communities.

These findings suggest that while azithromycin MDA is strongly associated with population density (as expected), the relationship between population density and MLS resistance is more modest, indicating that factors beyond population density influence resistance dynamics.

Population density estimates were sourced from the High-Resolution Settlement Layer by Data for Good at Meta (accessed at https://data.humdata.org/dataset/highresolutionpopulationdensitymaps-ner on 13 November 2024). Figure created using script https://osf.io/v4p5e.

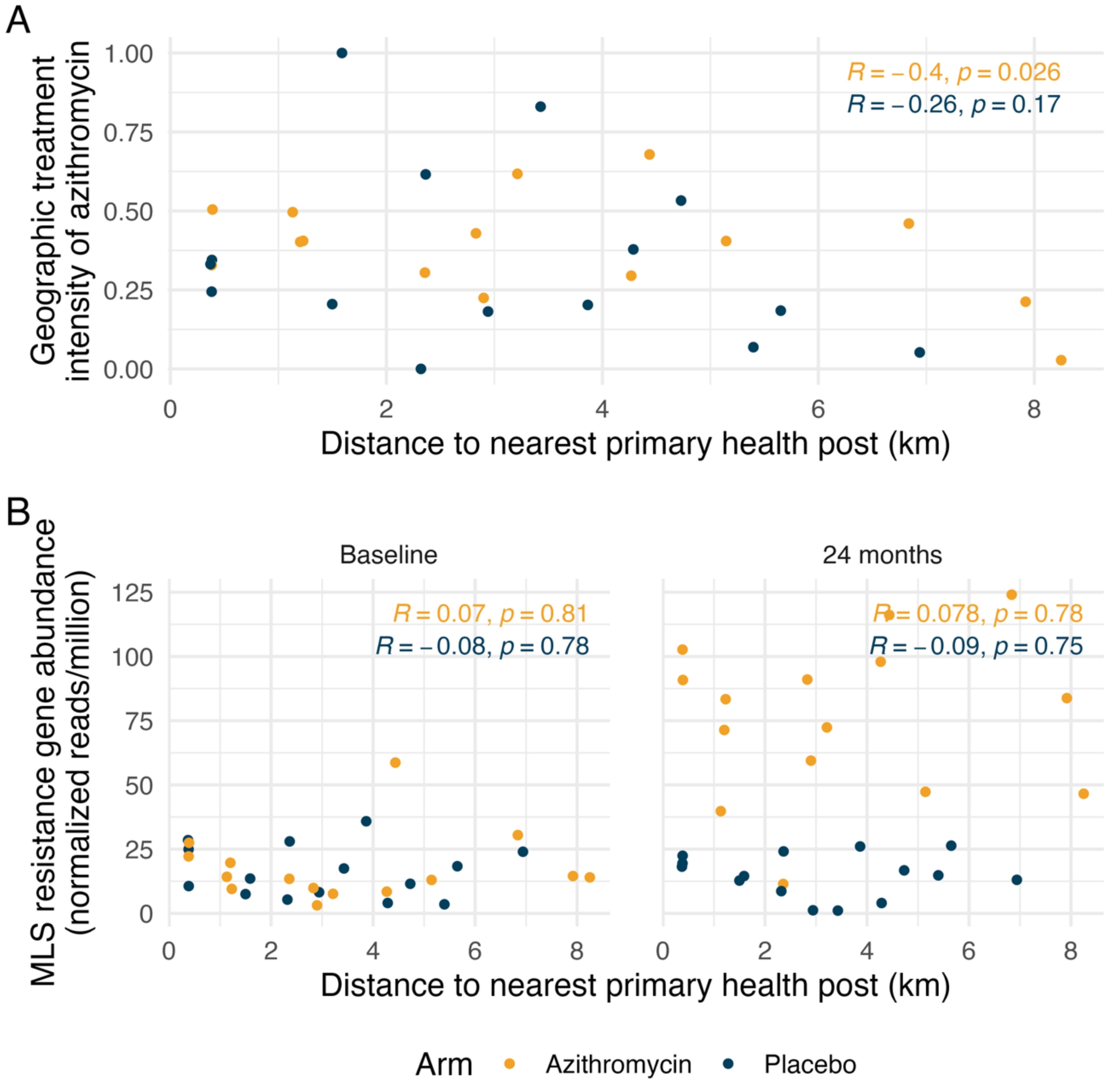


**Supplementary Figure 5: Relationship between geographic treatment intensity of azithromycin and subsequent MLS resistance by distance to the nearest primary health post**

(A) Relationship between distance to the nearest primary health post and the geographic treatment intensity of azithromycin at AMR monitoring villages, showing weak and non-significant associations in both, azithromycin- and placebo-treated villages.

(B) Correlation between distance to the nearest primary health post and MLS resistance at baseline (left) and 24 months (right). No significant associations are observed at either time point, suggesting that proximity to health posts does not strongly influence MLS resistance levels. These findings indicate that distance to primary health facilities is not a major confounder in the relationship between azithromycin MDA and antimicrobial resistance in this setting. Figure created using script https://osf.io/v4p5e.

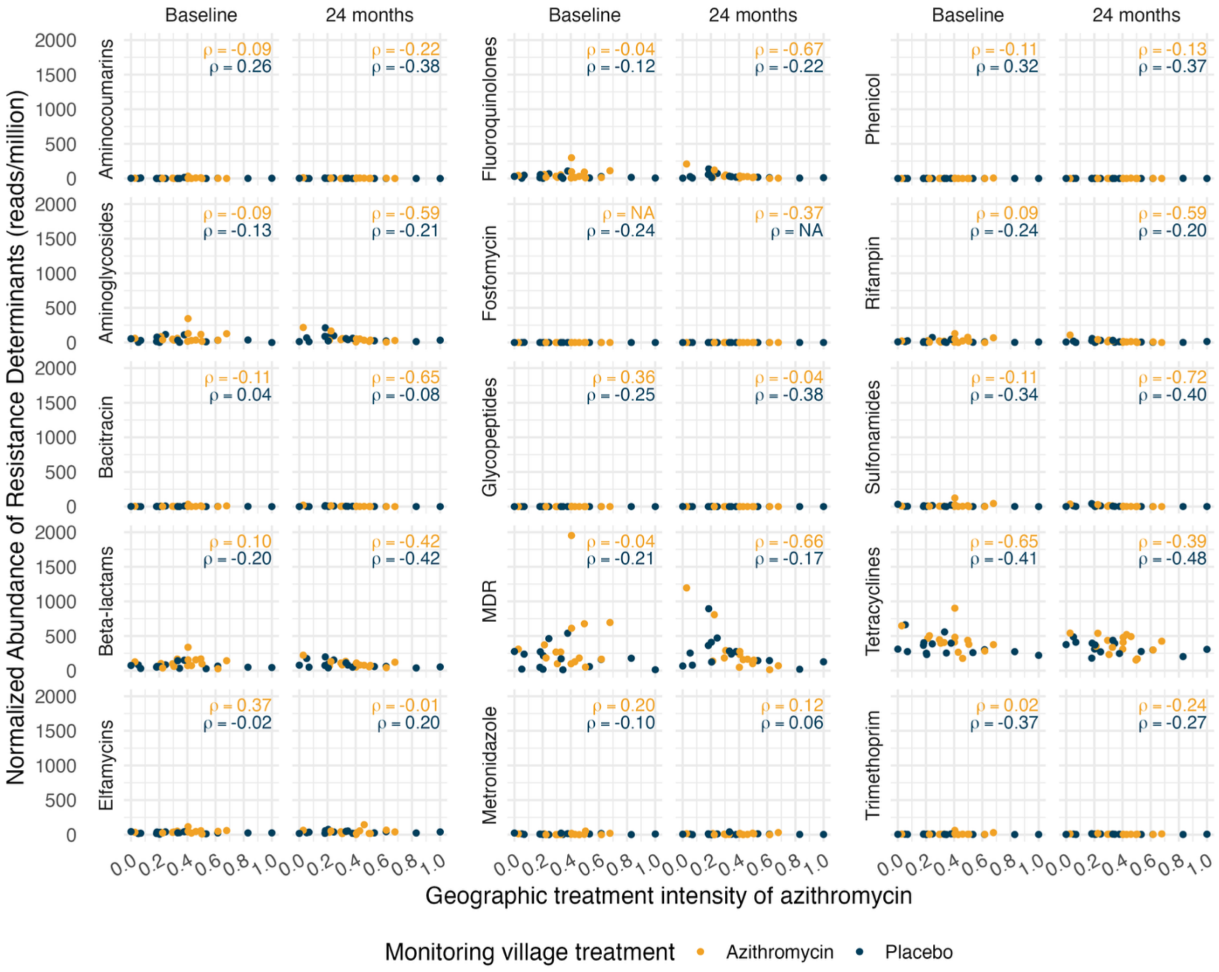


**Supplementary Figure 6: Relationship between azithromycin treatment intensity and the normalized abundance of resistance determinants across multiple antibiotic classes at baseline and 24 months.**

The scatterplots depict the normalized abundance of resistance determinants (reads per million) across 15 antibiotic classes in 30 AMR monitoring villages. For each antibiotic class, results are shown at baseline (left column of each pair) and at 24 months (right column of each pair), grouped by treatment arm: azithromycin-treated villages (yellow points) and placebo-treated villages (blue points). For each subplot, Spearman's correlation coefficient (ρ) is displayed, quantifying the association between treatment intensity and resistance abundance.

Overall, no consistent associations are observed between azithromycin treatment intensity and resistance determinants across antibiotic classes in either treatment arm, demonstrating no evidence of geographic spillover effects on AMR across a range of antibiotic classes and multidrug resistance (MDR).
Figure created using script https://osf.io/v4p5e.

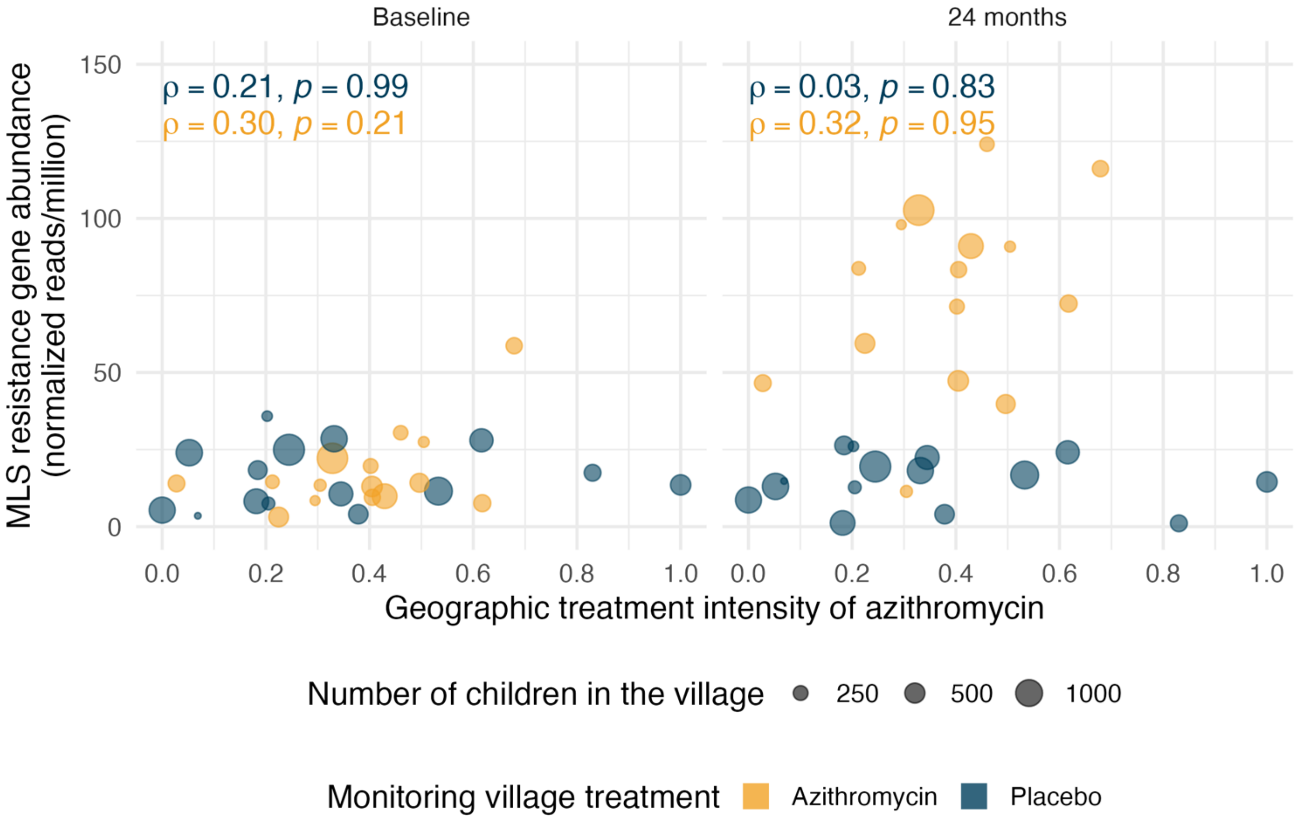


**Supplementary Figure 7: Relationship between geographic treatment intensity of azithromycin and MLS resistance gene abundance at baseline and 24 months by the number of children in the village**. Each point represents an AMR monitoring village, with point size corresponding to the number of children aged 1–59 months residing in the village, a proxy for village size. Colors indicate whether the AMR monitoring village received azithromycin (orange) or placebo (blue). Spearman correlation coefficients (ρ) and p-values are shown for each treatment group. There is no clear association between monitoring village size and either treatment intensity or observed resistance levels. Figure created using script https://osf.io/v4p5e.
